# Time to Discharge and Associated Factors Among Preterm Neonates Admitted to Kiwoko Hospital, Nakaseke District, Uganda: A Competing Risks Analysis

**DOI:** 10.64898/2026.04.13.26350793

**Authors:** Mutibwa Solomon, Wandiembe Symon, Mbonye Kayitale, Dick Nsimbe

## Abstract

**Background:** Preterm births contribute to approximately 35% of neonatal deaths globally, with an estimated 13.4 million infants born prematurely each year. Despite this substantial burden, limited evidence exists on time to discharge and its determinants among preterm neonates admitted to Neonatal Intensive Care Units (NICUs), particularly in rural Ugandan settings. This study aimed to investigate time to discharge and associated factors among preterm neonates admitted to Kiwoko Hospital in Nakaseke District, Uganda.

**Methods:** A retrospective cohort study was conducted using secondary data from Kiwoko Hospital on preterm neonates admitted to the Neonatal Intensive Care Unit (NICU) between 2020 and 2021 (n = 847). The cumulative incidence function was used to estimate the probability of discharge within 28 days of admission, accounting for competing events. A Fine and Gray sub-distribution hazard regression model was fitted to identify factors associated with time to discharge.

**Results:** Of the 847 preterm admissions, 70.1% were discharged alive within 28 days. The median time to discharge was 14 days. The cumulative incidence of discharge by 28 days was 68%, accounting for competing events. During follow-up, 165 neonates did not complete the 28-day period, including 88 deaths. Factors significantly associated with time to discharge included place of delivery (SHR: 0.62; 95% CI: 0.53–0.73; p<0.001), maternal residence in other districts (SHR: 0.69; 95% CI: 0.48–0.99; p=0.044), extreme preterm (SHR: 0.05; 95% CI: 0.03–0.09; p<0.001), very preterm (SHR: 0.18; 95% CI: 0.14–0.25; p<0.001), moderate preterm (SHR: 0.59; 95% CI: 0.46–0.76; p<0.001), triplet births (SHR: 0.40; 95% CI: 0.23–0.68; p=0.001), 2–4 ANC visits (SHR: 0.70; 95% CI: 0.56– 0.87; p=0.002), ≤1 ANC visit (SHR: 0.64; 95% CI: 0.49–0.85; p=0.002), respiratory distress syndrome (SHR: 0.64; 95% CI: 0.48–0.74; p<0.001), and birth trauma (SHR: 2.62; 95% CI: 1.60– 4.29; p<0.001).

**Conclusions:** Respiratory distress syndrome, fewer antenatal care visits, out-of-district residence, and higher degrees of prematurity were associated with prolonged time to discharge among preterm neonates. Strengthening antenatal care utilization and improving access to quality neonatal care in underserved areas may enhance discharge outcomes.

## Background

Preterm births are defined as deliveries that occur before 37 completed weeks of gestation (1).The World Health Organization (WHO) categorizes preterm births into three subgroups based on gestational age: extremely preterm (less than 28 weeks), very preterm (28 to 32 weeks), and moderate to late preterm (32 to 37 weeks) (2). Preterm births may occur spontaneously or result from medical interventions due to complications such as infections or other pregnancy-related issues that necessitate early induction or cesarean delivery (1,2). In 2020, over 13.4 million babies were born prematurely worldwide, with 65% of these births occurring in sub-Saharan Africa and South Asia regions that accounted for approximately 3.9 million and 4.8 million preterm births, respectively (2,3). Preterm birth is associated with numerous complications, including immature lungs, difficulty in regulating body temperature, infections, feeding difficulties, and poor weight gain. These complications significantly contribute to neonatal mortality, making preterm birth the second leading cause of infant mortality globally (4). Sub-Saharan Africa and South Asia bear the heaviest burden of neonatal mortality, with preterm births accounting for a significant proportion of these deaths. Approximately 1.1 million neonatal deaths annually are attributed to prematurity, representing a considerable portion of the 2.4 million neonatal deaths worldwide (2).

In East Africa, neonatal mortality is estimated to be approximately 24.2% of the global rate, with prematurity contributing to 52% of this figure (5). In Uganda, neonatal mortality is reported at 19.2 per 1000 live births, resulting in approximately 110,000 deaths annually, with complications related to prematurity responsible for around 30% of these deaths (6–8). These statistics underscore the urgent need for targeted research and interventions aimed at reducing neonatal mortality rates among preterm neonates and improving the quality of their care. Numerous studies have been conducted globally, as well as in East Africa, Kenya, and Uganda, to investigate the incidence, prevalence, and impact of preterm births, with a particular emphasis on neonatal mortality. For example, an East African study revealed that 52% of neonatal deaths were attributed to preterm complications (5). In Kenya, a study found that 78.4% of neonatal deaths occurred within the first week of life, primarily due to preterm complications (9). Furthermore, research conducted at a rural NICU showed that 14% of neonatal deaths were linked to preterm birth (10). A more recent study in Uganda between 2015 and 2019 reported an incidence of 13 preterm births per 1000 births (11).

Preterm neonates often require specialized care in the NICU to address complications such as temperature regulation, respiratory difficulties, and feeding challenges while their vital organs, including the brain, lungs, and liver, mature (12,13). Several studies have been conducted to explore strategies for reducing preterm-related complications within the NICU, aiming to decrease preterm neonatal mortality and improve overall neonatal outcomes. Some of the key studies include a retrospective study by Gebreheat and Teame (14) on survival and mortality of preterm neonates in a NICU in Northern Ethiopia, as well as studies by Yismaw et al. (15), Lin et al. (16), Bereka et al. (17), Egesa et al. (18), and Opio et al. (19) in various African contexts, all of which focused on survival and mortality predictors in preterm neonates. While these studies have provided valuable insights into the predictors of survival and neonatal mortality, they have largely overlooked another crucial aspect of preterm neonatal care: the time to discharge from the NICU. Understanding the duration of NICU stays and the factors influencing early or prolonged discharges is vital for effective resource planning, healthcare management, and ensuring positive neonatal outcomes. Prolonged NICU stays increase the likelihood of complications, such as nosocomial infections and medication side effects, which not only impose significant financial burdens on families but also strain healthcare systems, thereby increasing the risk of mortality (20). Conversely, earlier discharge can reduce the risks of infection and adverse medication effects, while also improving treatment quality, ultimately contributing to better survival outcomes and overall neonatal health (21,22).

Furthermore, A recent systematic review by Fu et al. (23) examined the risk factors associated with length of stay and time to discharge in the NICU. The review identified key factors such as birth weight, gestational age, sepsis, necrotizing enterocolitis, bronchopulmonary dysplasia, and retinopathy of prematurity as significant predictors of NICU stay. However, it also highlighted the scarcity of high-quality studies on this subject and emphasized the need for more well-designed research. additionally, the existing literature has largely failed to consider competing risks, such as death, which may influence the time to discharge and the interpretation of length of stay data. It’s against this background that this study aims to fill the critical gap in the literature, including the limited research on time to discharge and its associated factors, the lack of consideration of death as a competing risk. By focusing on the temporal aspects of preterm neonatal care, this research seeks to provide valuable insights that can inform evidence-based interventions, optimize NICU practices, and reduce unnecessary delays in discharge. Ultimately, this research aims to minimize preterm neonatal mortality, contribute to better neonatal outcomes, and support the achievement of Sustainable Development Goal (SDG) 3.2, which targets the reduction of neonatal mortality.

## Methods

The study was conducted using secondary data from the Neonatal Intensive Care Unit (NICU) at Kiwoko Hospital in central Uganda, a private, not-for-profit facility serving as a referral center for three districts (10). Nakaseke, Luwero, and Nakasongola, which collectively have a population of approximately 1,000,000 people. The NICU, which has been operational since 2001, specializes in the care of preterm and critically ill newborns and operates 38 beds, admitting over 1,200 infants annually. This retrospective cohort study analyzed data from preterm neonates admitted between 2020 and 2021, tracking outcomes over 28 days post-admission. The data extraction process involved reviewing patient charts using a Google Forms based tool, and a final sample of 847 chart files was included after excluding incomplete records.

### Measures of Outcome

The dependent variable in this study was time to discharge, or length of stay, measured in days. This was categorized into three outcomes: (1) earlier discharge time alive within 28 days, (2) earlier discharge time within 28 days due to death, and (3) prolonged discharge time beyond 28 days. The primary outcome of interest was earlier time to discharge alive (within 28 days), while prolonged discharge time (beyond 28 days) was treated as a censored outcome. Discharge outcomes due to death were considered competing events, as they contradicted the primary outcome of earlier discharge alive. According to Fine and Gray (26), the competing event (death) prevents the occurrence of the primary outcome (earlier discharge alive) permanently, as the primary outcome is no longer possible once death occurs. Therefore, death was treated as a competing event in the analysis.

### Measures of explanatory variables

The independent variables included maternal socio-economic factors, obstetrical factors, and preterm characteristics obtained from chart files of preterm neonates within the NICU at Kiwoko Hospital. Maternal socio-economic factors included maternal residence (Luwero, Nakaseke, Nakasongola, and others), mode of transport (ambulance and public means), and maternal age (14 to 45 years). Obstetrical factors included number of antenatal visits (≤1, 2-4, and ≥5), place of delivery (inbound and outbound), mode of delivery (C-section and vaginal), and maternal HIV status (negative, positive, and unknown). Preterm characteristics included preterm weight group (ELBW, VLBW, and LBW), preterm hypothermia status (no and yes), preterm neonatal sepsis (no and yes), preterm birth trauma status (no and yes), KMC use (no and yes), preterm gestation group (extreme preterm, late preterm, moderate preterm, and very preterm), and gestational age (21 to 39 weeks).

### Statistical Analysis

The analysis was performed using STATA software, version 14, at three levels: univariate, bivariate, and multivariate. At the univariate level, data summaries were generated using proportions and frequencies. The time to earlier discharge, with respect to death as a competing event, was estimated using the cumulative incidence estimator. Cumulative incidence curves were employed to summarize the probabilities of discharge within 28 days of admission to the NICU. As per Gooley et al. (27), the Kaplan-Meier estimator is biased when competing events are treated as censored, assuming that the individual could still experience the event of interest, which is not feasible. Therefore, the Kaplan-Meier estimator was rejected in favor of the cumulative incidence estimator. To identify the potential variables associated with time to discharge, these were tested at bivariate level using log-rank test for equality of survival (with a significance level set at p < 0.25 for categorical variables) and cox test for equality of survival for count and continuous variables. The study considered 0.25 level of significance so as to identify more of the potential factors associated with time to discharge and as well keeping in mind existence of more refining tests through regression diagnostics and multivariate level analysis (28–30). At multivariable level, a Sub-distribution hazard regression model of Competing risk events to investigate the association between time to discharge and the identified covariates was used. The study adopted this model so as to consider the competing event of earlier discharge time due to death while estimating the effect of the identified covariates on the sub-distribution hazard rate (31,32). The study adopted the Gray’s diagnostic test for proportional sub-distribution hazards to confirm that the relative sub-hazard is fixed over time (26,33). The proportional sub-hazards assumption was considered violated (null hypothesis rejected with p < 0.05) when time interactions with a given potential predictor were significant.

## Results

**Table** 1 presents the distribution of preterm neonates categorized by maternal socio-economic, obstetric, and preterm-related characteristics. It also summarizes the time to discharge for the preterm neonates, revealing that 70.1% achieved the primary outcome of earlier discharge. However, 10.4% of neonates died during the 28-day follow-up period, and 19.5% were censored due to prolonged hospitalization, remaining in the NICU at the end of the 28-day period. The analysis further highlights that the age of the youngest mother was 14 years, while the oldest mother was 45 years. The gestational age of the neonates ranged from a minimum of 21 weeks to a maximum of 39 weeks. The number of ANC visits varied from a minimum of 0 visits to a maximum of 9 visits. The lowest APGAR score recorded for a preterm neonate upon admission was 1, while the highest at 5 minutes was 10. The majority of mothers of preterm neonates were identified as married (84.1%), adults (83.7%), residing in Luwero district (48.5%), and relied on public transportation (75.7%) to reach the hospital. 64.9% of the mothers attended between 2 and 4 ANC visits, 52.2% delivered within the hospital of preterm admission (inbound), and 71.2% delivered by normal birth (vaginal). Most mothers (65.2%) offered KMC practice to their babies during admission. 51.1% of the preterm neonates were male, 67.9% were singletons, and 64.9% had low birth weight. The preterm neonates that survived the neonatal period were 759, while 88 were lost due to death, with apnea being the main cause of death. A log-rank test was carried out to examine the association between time to discharge and the different explanatory variables. The analysis revealed that significant variables at the 75% confidence level included maternal residence, place of delivery, mode of delivery, number of gestations, maternal HIV status, preterm neonatal sepsis, preterm birth trauma, KMC use, preterm gestational group, ANC visits, preterm weight group, preterm admission age, APGAR score at 1 minute, and APGAR score at 5 minutes.

**Table 1.** Differential in time to discharge by selected socio-economic, obstetrics and preterm related Factors.

| Covariates | N | Percentage | Median time to discharge in days | Log-rank chi2 ($\chi^2$), p |
| --- | --- | --- | --- | --- |
| **Discharge** |  |  |  |  |
| Earlier discharge | 594 | 70.1 |  |  |
| Discharge due to death | 88 | 10.4 |  |  |
| Prolonged discharge | 165 | 19.5 |  |  |
| **Maternal Residence** |  |  |  |  |
| Luwero | 411 | 48.5 | 12 |  |
| Nakaseke | 237 | 27.9 | 16 |  |
| Nakasongola | 137 | 16.2 | 13 | 11.42 |
| Other | 62 | 7.3 | 24 | (p = 0.010) |
| **Mode of transport** |  |  |  |  |
| Ambulance | 206 | 24.3 | 16 | 3.55 |
| Public means | 641 | 75.7 | 13 | (p = 0.600) |
| **Number of antenatal Visits (times)** |  |  |  |  |
| ≤1 | 183 | 21.6 | 22 |  |
| 2-4 | 550 | 64.9 | 14 | 32.65 |
| ≥5 | 114 | 13.5 | 8 | (p = 0.001) |
| **Preterm weight group** |  |  |  |  |
| ELBW | 77 | 9.4 | 0 |  |
| VLBW | 209 | 25.6 | 0 | 348.82 |
| LBW | 530 | 65.0 | 8 | (p = 0.001) |
| **Place of delivery** |  |  |  |  |
| Inbound | 442 | 52.2 | 10 | 15.11 |
| Outbound | 405 | 47.8 | 16 | (p = 0.001) |
| **Mode of delivery** |  |  |  |  |
| C-section | 244 | 28.8 | 10 | 7.79 |
| Vaginal | 603 | 71.2 | 15 | (p = 0.005) |
| **Number of gestations** |  |  |  |  |
| Singleton | 575 | 67.9 | 14 |  |
| Triplet | 18 | 2.1 | 0 | 7.16 |
| Twin | 251 | 30.0 | 13 | (p = 0.028) |
| **Maternal syphilis status** |  |  |  |  |
| Negative | 666 | 78.6 | 13 |  |
| Positive | 45 | 5.3 | 11 | 5.88 |
| Unknown | 136 | 16.1 | 16 | (p = 0.053) |
| **Maternal HIV status** |  |  |  |  |
| Negative | 722 | 85.2 | 13 |  |
| Positive | 69 | 8.2 | 14 | 6.67 |
| Unknown | 56 | 6.6 | 25 | (p = 0.040) |
| **Maternal UTI status** |  |  |  |  |
| Negative | 367 | 43.3 | 15 |  |
| Positive | 143 | 16.9 | 10 | 8.75 |
| Unknown | 337 | 39.8 | 14 | (p = 0.013) |
| **Preterm hypothermia status** |  |  |  |  |
| No | 721 | 85.1 | 13 | 1.64 |
| Yes | 126 | 14.9 | 15 | (p = 0.201) |
| **Preterm neonatal sepsis** |  |  |  |  |
| No | 796 | 94.0 | 14 | 6.14 |
| Yes | 51 | 6.0 | 9 | (p = 0.013) |
| **Preterm RDS status** |  |  |  |  |
| No | 687 | 81.1 | 12 | 16.43 |
| Yes | 160 | 18.9 | 22 | (p < 0.001) |
| **Preterm birth trauma status** |  |  |  |  |
| No | 843 | 99.5 | 14 | 3.74 |
| Yes | 4 | 0.5 | 6 | (p = 0.053) |
| **KMC use** |  |  |  |  |
| No | 295 | 34.8 | 6 | 118.07 |
| Yes | 552 | 65.2 | 18 | (p < 0.001) |
| **Preterm gestational group** |  |  |  |  |
| Extreme Preterm | 102 | 12.0 | 0 |  |
| Late Preterm | 48 | 5.7 | 6 |  |
| Moderate Preterm | 436 | 51.5 | 8 | 299.57 |
| Very preterm | 261 | 30.8 | 27 | (p < 0.001) |
| **Overall** | **847** | **100** | **14** |  |

### Median time to discharge

**Table** 1 further shows that the overall median time to discharge was 14th day of follow-up after admission. By maternal residence, preterm neonates whose mothers resided in Luwero had the earliest median time to discharge, on the 12th day, while those whose mothers resided in other districts (excluding Nakaseke and Nakasongola) experienced the most delayed median discharge, on the 24th day. Regarding maternal ANC visits, preterm neonates whose mothers had more than five visits had the shortest median time to discharge, on the 8th day, followed by those whose mothers had between two and four visits, discharged on the 14th day. In contrast, neonates whose mothers had at most one visit had a median discharge time of 22 days. For preterm neonates grouped by number of gestations, twins had an earlier median time to discharge, on the 13th day, compared to singletons, whose median time was the 14th day. With respect to preterm gestational group, late preterm neonates had the earliest median discharge time on the 6th day, followed by moderate preterm neonates on the 8th day, while very preterm neonates experienced a delayed median discharge on the 27th day.

### Probability of discharge within 28 days

**Fig. 1** presents the cumulative incidence estimates for the probability of discharge within 28 days of admission. Overall, 68% of preterm neonates were discharged from the NICU within this period. Regarding place of residence, the probability of discharge within 28 days was lowest (54%) for preterm neonates whose mothers resided outside Nakaseke, Luweero, and Nakasongola districts. By preterm gestational age, the probability of discharge was very low (18%) among extreme preterm neonates compared to very preterm (48%), moderate preterm (87.7%), and late preterm (97%) neonates. By place of delivery, preterm neonates born outside the hospital of admission had a lower probability of discharge (59%) than those born inbound (76%). The probability of being discharged within 28 days was lowest among triplets (40%), compared to twins (67.5%) and singletons (72%). According to RDS status, those diagnosed with RDS had a lower likelihood of discharge (53%) compared to those without RDS (71.6%). Interestingly, preterm neonates with birth trauma had a higher probability of discharge (95%) than those without (68%). Finally, based on maternal ANC visits, neonates whose mothers attended five or more visits had a higher probability of discharge (80%) than those whose mothers attended fewer visits.

**Fig. 1.**
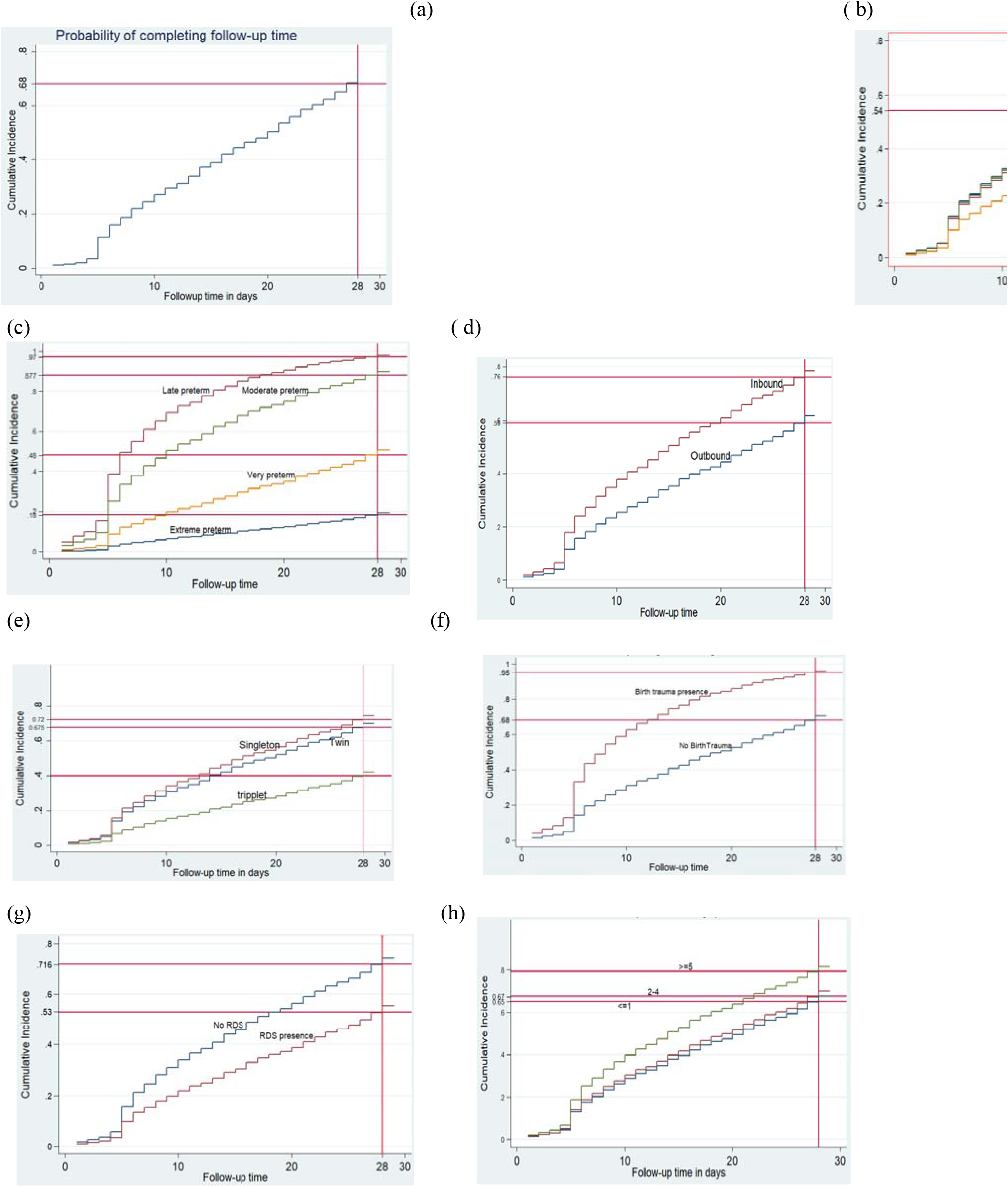
Cumulative incidence curve estimates showing the probability of discharge within 28 days by selected socio-economic, obstetrics and preterm related factors: **(a)** Overall discharge probability, **(b)** Maternal residence, **(c)** Preterm gestation group, **(d)** Place of delivery, **(e)** Number of gestations, **(f)** Birth Trauma Status, **(g)** RDS Status, **(h)** Number of ANC Visits

### Time to discharge and associated Factors

At this level, all variables that were identified as statistically significant at bivariate level and time invariant, were included in the general sub-hazard regression model to analyze their association with time to discharge as shown in the **Table** 2. Among the maternal residences, other places of residences depicted a significant statistical association with time to discharge among preterm neonates in the NICU (p = 0.044). The corresponding sub-hazard ratio 0.69 implied that preterm neonates whose mothers were from other places of residence had a 31% lower likelihood of having an earlier discharge from the NICU than those whose mothers were from Nakaseke district. Hence, the longer the distances the preterm mothers travel, the lower the likelihood of early discharge by their preterm babies. Preterm neonates delivered from outbound/ out of Nakaseke hospital showed a significant association (p< 0.001) with delayed time to discharge. The sub-hazard ratio 0.62 implied that preterm neonates born from outside the main hospital were 38% less likely to have an earlier discharge than those who were born from within the hospital of admission. Regarding the number of gestations, the triplet preterm neonates exhibited a significant statistical association with time to discharge (p< 0.001). They showed 60% triplets (SHR: 0.40; 95% CI: 0.23-0.68; p = 0.001) less likelihood of having an earlier discharge from the hospital NICU as compared to the singleton neonates. With the preterm gestational age groups, the extreme preterm neonates were 95% (SHR: 0.62; 95% CI: 0.53-0.73; p < 0.001) less likely to have an early discharge as compared to the late preterm, followed the very preterm with 82% (SHR: 0.18; 95% CI: 0.14, 0.25; p<0.001) less likelihood and the moderate with 41% less likelihood (SHR: 0.59; 95% CI: 0.46-0.76; p<0.001). This meant that there is a higher likelihood of earlier discharge as gestational ages near term and vice versa. These results were statistically evidenced by the respective p< 0.001. Preterm neonates admitted with RDS depicted a significant statistical effect on delayed time to discharge (p< 0.001). The corresponding SHR of 0.64 implied that preterm neonates diagnosed with RDS were 36% less likely to have an early discharge from the NICU than those without RDS. Fortunately, preterm neonates diagnosed with a birth trauma were evidenced to be 2.6 times (SHR: 2.62; 95% CI: 1.60, 4.29; p<0.001) more likely to have an early discharge than those without the trauma (p< 0.001). In regards to number of maternal antenatal care visits, preterm neonates whose mothers attended between 2 and 4 antenatal visits, were 30% less likely to have an earlier discharge from the NICU as compared to those whose mothers attended 5 and above. For those whose mothers attended at most one, they had a 36% less likelihood of leaving the NICU earlier as compared to those whose mothers had 5 and above (p-values: 0.002 & 0.002 respectively). The general multivariate model was tested for proportional Sub-hazards assumption and variables whose p-value were significant implied that their relative Sub hazards or coefficients were time varying hence violating the assumption. The p-values of KMC use (p< 0.001), mode of transport (.028) and weight group (0.001) implied that they were time varying hence violating the proportional sub-hazard assumption. These were therefore eliminated for multivariate analysis.

**Table 2.** Final Sub-hazard regression model linking the time to discharge to socio-economic, obstetrics and preterm related Factors

| Covariates | Sub-hazard Ratio | P-value | 95% CI |
| --- | --- | --- | --- |
| **Maternal Residence** |  |  |  |
| Nakaseke (*Ref*) | - | - | - |
| Luwero | 1.07 | 0.508 | 0.88, 1.29 |
| Nakasongola | 1.04 | 0.776 | 0.80, 1.34 |
| Other | 0.69 | **0.044** | 0.48, 0.99 |
| **Place of delivery** |  |  |  |
| Inbound (*Ref*) | - | - | - |
| Outbound | 0.62 | **<0.001** | 0.53, 0.73 |
| **Number of gestations** |  |  |  |
| Singleton (*Ref*) | - | - | - |
| Triplet | 0.40 | **0.001** | 0.23, 0.68 |
| Twin | 0.88 | **0.157** | 0.75, 1.05 |
| **Preterm gestational age group** |  |  |  |
| Late preterm (*Ref*) | - | - | - |
| Extreme preterm | 0.05 | **<0.001** | 0.32, 0.93 |
| Moderate Preterm | 0.59 | **<0.001** | 0.46, 0.76 |
| Very Preterm | 0.18 | **<0.001** | 0.14, 0.25 |
| **Preterm RDS status** |  |  |  |
| No (*Ref*) | - | - | - |
| Yes | 0.60 | **<0.001** | 0.48, 0.74 |
| **Preterm birth trauma status** |  |  |  |
| No (*Ref*) | - | - | - |
| Yes | 2.62 | **<0.001** | 1.60, 4.29 |
| **Number of antenatal visits (times)** |  |  |  |
| ≥5 (*Ref*) | - | - | - |
| 2-4 | 0.70 | **0.002** | 0.56, 0.87 |
| ≤1 | 0.64 | **0.002** | 0.49, 0.85 |

*Ref.* Reference category; *CI* – confidence interval

## Discussion

This study aimed to examine the time to discharge and associated factors among preterm neonates admitted to the NICU at Kiwoko Hospital, using a competing risks approach over a 28-day follow-up period. Place of delivery, preterm gestational age group, number of gestations, number of antenatal care (ANC) visits, maternal residence, preterm respiratory distress syndrome (RDS) status, and trauma status were identified as significant factors influencing the time to discharge. Specifically, preterm neonates whose mothers resided outside Nakasongola and Luwero districts were less likely to be discharged early. One plausible explanation is that longer travel distances to the hospital may delay access to timely prenatal and postnatal care, resulting in poorer maternal and neonatal health outcomes that prolong hospitalization. Although no prior studies were found that directly linked maternal district of residence to neonatal discharge timing, existing literature supports the broader relationship between geographical access and neonatal outcomes. For example, a study in Wales demonstrated that for every 15-minute increase in travel time to a neonatal intensive care unit (NICU), the odds of early neonatal death increased significantly (34). Similarly, research from the United States indicated that longer driving distances to perinatal care centres were associated with higher NICU admission rates and adverse neonatal outcomes (35). A systematic review by Nesbitt et al. (36) further revealed that increased distance to health facilities significantly reduced the utilization of maternal and new-born health services in low- and middle-income countries. Qualitative evidence also suggests that families residing far from NICUs face challenges such as limited visitation and reduced parental involvement in care, which can contribute to longer hospitalization durations (37). Collectively, these findings lend support to the observed association between maternal residence and delayed neonatal discharge.

Regarding gestational age, neonates classified as extreme, very, and moderate preterm experienced a significantly longer time to discharge compared to those born late preterm. This finding aligns with existing literature, which consistently demonstrates that the degree of prematurity is a key determinant of neonatal outcomes and length of hospital stay. Preterm infants, particularly those born before 32 weeks of gestation, often require extended hospitalization due to complications such as respiratory distress syndrome, feeding difficulties, temperature instability, and heightened susceptibility to infections (38,39). Furthermore, studies have shown that as gestational age decreases, the likelihood of requiring prolonged intensive care increases substantially, thereby delaying discharge (40,41).

With regard to the number of gestations, triplet preterm neonates were significantly more likely to experience delayed discharge compared to singletons. This finding is consistent with existing literature indicating that multiple births, particularly triplets and higher-order multiples, are associated with prolonged neonatal intensive care unit (NICU) stays. Finnbogadóttir et al. (42) reported that triplet pregnancies often result in earlier deliveries, low birth weights, and a higher incidence of neonatal complications, all of which contribute to extended hospitalizations. Similarly, Whyte et al. (43) observed that the risk of respiratory and metabolic complications increases with multiple births, which in turn lengthens the time to discharge. Verberg et al. (44) also noted that multiple gestations are inherently high-risk due to factors such as uterine crowding and placental insufficiency, increasing the likelihood of adverse neonatal outcomes and the need for intensive medical care.

The number of maternal antenatal care (ANC) visits was another significant factor. Specifically, neonates born to mothers with one or no ANC visits were less likely to be discharged early compared to those whose mothers attended five or more visits. This underscores the critical role of consistent prenatal care in improving neonatal outcomes. ANC visits provide essential opportunities for the early detection and management of pregnancy-related complications, appropriate monitoring of fetal development, and timely health education for expectant mothers, all of which contribute to better perinatal and neonatal outcomes (45,46). Several studies have demonstrated that inadequate ANC is associated with an increased risk of preterm birth, low birth weight, and neonatal morbidity, which can prolong hospital stays (47,48).

Preterm neonates born outside the hospital (outborn) were less likely to be discharged early compared to their inborn counterparts. This is due to delays in accessing timely neonatal interventions or unmanaged complications at the time of birth, which can negatively affect early stabilization and recovery (49). Regarding birth trauma, interestingly, neonates without recorded trauma experienced delayed discharge compared to those with trauma. One possible explanation is that the trauma cases may have been minor and promptly addressed, whereas the non-trauma group may have included neonates with undetected or unrecorded complications that required longer care, as discussed by (50). Additionally, the diagnosis of respiratory distress syndrome (RDS) was significantly associated with prolonged hospital stays. RDS, primarily caused by surfactant deficiency in preterm neonates, necessitates specialized respiratory support and intensive monitoring, thereby delaying discharge (51). This finding aligns with the results of Egesa et al. (18), who identified RDS as a major contributor to extended NICU admissions among preterm infants. In contrast, other variables such as mode of delivery and APGAR scores were not significantly associated with time to discharge in the current study, suggesting that early neonatal outcomes may be more strongly influenced by underlying morbidity than by these perinatal indicators.

In summary, this study utilized secondary data of 847 preterm neonates admitted to Kiwoko Hospital NICU and followed for 28 days. Of these, 549 were discharged alive within the neonatal period, 165 were still admitted at the end of follow-up, and 88 died. Time to death was treated as a competing risk, and analysis was conducted using a Cumulative Incidence Estimator and the Sub-distribution Hazard Model to account for these competing events. The median time to discharge was 14 days. Significant factors associated with discharge included gestational age group, triplet births, trauma status, and RDS status among the neonates, as well as maternal place of delivery, ANC attendance, and district of residence. Conversely, APGAR scores and mode of delivery showed no significant effect on discharge timing.

There are some limitations that need to be put into consideration. The use of secondary data limited the opportunity for direct interaction with respondents, and missing information in some preterm neonatal chart files may have affected the reliability and validity of the findings (52). Future research would benefit from prospective study designs and the incorporation of qualitative components to provide a more comprehensive understanding of the factors influencing time to discharge among preterm neonates.”

## Conclusion

The study revealed that the median time to discharge was 14 days. Preterm neonates diagnosed with respiratory distress syndrome (RDS), those born to mothers with fewer than five antenatal care (ANC) visits, those delivered outside hospitals equipped with NICUs, and those whose mothers resided in distant districts were less likely to be discharged early. This suggests that increasing ANC attendance, improving the diagnosis and management of neonatal complications, and enhancing NICU accessibility could significantly improve discharge outcomes. In turn, this could reduce preterm neonatal mortality and support progress toward achieving Sustainable Development Goal (SDG) 3.2, which aims to lower neonatal mortality to 12 per 1,000 live births. Healthcare providers should prioritize sensitizing expectant mothers on the importance of consistent ANC attendance, as this enables early identification and management of complications. The government should ensure adequate staffing of NICUs by deploying sufficient medical personnel to offer timely and continuous care for preterm neonates with emerging complications such as RDS and birth trauma. Moreover, healthcare professionals should develop individualized care strategies for singletons, twins, triplets, and higher-order multiples, addressing their unique developmental and medical needs. Government investment is also needed to expand NICU services to more health facilities across regions, which would help reduce travel distances, minimize delays in care, and promote equity in neonatal health outcomes. Future research should incorporate primary data collection from mothers and healthcare workers to gain deeper insights into the care processes and barriers to early discharge. Including additional clinical indicators not captured in admission records and exploring alternative outcomes such as readmissions or new complications post-discharge could enhance understanding of prolonged NICU stays. Additionally, conducting similar studies across hospitals with different NICU capacities could uncover regional disparities in discharge predictors and inform targeted interventions.

## Data Availability

The data used in this study were obtained from Kiwoko Hospital and are not publicly available due to institutional confidentiality policies. Access to the data can be requested from the corresponding author, subject to approval by the hospital and relevant ethical authorities.

## Declarations

## Abbreviations

NICUs: Neonatal Intensive Care Units
NICU: Neonatal Intensive Care Unit
SHR: Sub-hazard Ratio
RDS: Preterm respiratory distress syndrome
WHO: World Health Organisation
HIV: Human Immunodeficiency Virus
ELBW: Extremely Low Birth Weight
VLBW: Very Low Birth Weight
LBW: Low Birth Weight
KMC: Kangaroo Mother Care
APGAR: Appearance, Pulse, Grimace, Activity, and Respiration
Ref: Reference Category
CI: Confidence Interval
ANC: Number of antenatal care
ANC: Maternal antenatal care
SDG: Sustainable Development Goal

## Acknowledgements

The authors extend their sincere gratitude to the management of Kiwoko Hospital for granting access to the data required for this study. Special thanks go to the clinical director and NICU staff for their cooperation and support during the data collection process.

## Funding

This study was self-funded. No external funding was received for the conduct of this research.

## Ethical Approval and Consent to Participate

The study utilized secondary data following approval from the Ethics Committee of the School of Statistics and Planning at Makerere University. Permission to access data was granted by the Clinical Director of Kiwoko Hospital. The research protocols, including data collection methods, participant involvement, and ethical considerations, were reviewed and approved by the relevant institutional and/or licensing committees during the original data collection. Informed consent was obtained from all participants or their legal guardians as part of the initial data collection process. Given that this study involved secondary data, the requirement for additional informed consent was waived by the Ethics Committee of the School of Statistics and Planning at Makerere University, in line with national regulations. The study adhered to the ethical and consent procedures set by Kiwoko Hospital. All procedures followed in this research complied with the ethical standards established by relevant national and institutional committees for human research, as well as the Declaration of Helsinki (1975, amended in 2008). Strict confidentiality measures were implemented to protect patient information.

## Consent for Publication

Not applicable.

## Clinical Trial Number

Not applicable

## Competing Interests

The authors declare no competing interests.

## Notes

### Competing Interest Statement

The authors have declared no competing interest.

### Funding Statement

This study did not receive any funding

